# Simulating population compliance with pandemic interventions using large language models

**DOI:** 10.64898/2026.05.12.26352942

**Authors:** Runzhou Liu, Claire Jong, Haoyang Li, Yiming Cao, Qing Yao, Teresa Yamana, Sen Pei, Hongru Du

## Abstract

Effective pandemic response requires accurate modeling of population compliance with non-pharmaceutical interventions (NPIs), yet most epidemic models treat behavioral change as fixed scenarios rather than an emergent process. Here, we test whether large language model (LLM)-based agents can generate individualized behavioral responses to time-varying NPIs and disease risk. We instantiate demographically representative agents in three U.S. cities (Boston, Denver, San Antonio) and condition them on evolving outbreak conditions and policies during the early COVID-19 pandemic, without fitting to observed mobility data. Across three frontier LLMs and their ensemble, agents generate zero-shot mobility changes across restaurants, retail, and entertainment venues, benchmarked against cellphone-derived foot-traffic records. The simulations recover average mobility trends across cities and venue types but exhibit overly narrow within-city variation. The three LLMs display distinct biases, while an ensemble approach improves robustness and overall performance. These findings establish LLM agents as a promising framework for modeling adherence to NPIs and highlight the need for further fine-tuning and empirical validation before they can support policy analysis.

## Introduction

Emerging infectious diseases pose continual threats to public health, economies, and human societies. In the early stages of outbreaks, non-pharmaceutical interventions (NPIs) are often the only available tools to suppress transmission and prevent healthcare system overload. Their success, however, depends on behavioral responses that vary across social, economic, and cultural contexts^1^. Despite this central role, policy adherence remains poorly captured in current epidemic models, which typically rely on fixed, scenario-based assumptions on population behavior change^2^. This simplification obscures critical heterogeneity in human behavior and limits models’ ability to inform effective interventions.

Cellphone-derived foot-traffic records provide a measurable and interpretable data source for quantifying population-level policy compliance^3^. These data track aggregated daily visits from communities to specific points of interest (POIs), such as restaurants, grocery stores, and workplaces, revealing how individuals adjust their routine activities during an outbreak^4^. As many NPIs aim to reduce contact in specific settings, location-resolved mobility patterns provide a direct behavioral signal of policy adherence. While prior studies have leveraged foot-traffic data to support high-resolution epidemic modeling and forecasting^5,6^, existing approaches largely treat compliance as an outcome to be fitted, rather than as a generative process arising from individual decision-making.

Large language models (LLMs) offer a new approach to modeling human behavior change as an emergent process grounded in individual decision-making. By encoding rich social knowledge and contextual reasoning, LLM-based agents can generate individualized behavioral responses conditioned on policy, risk perception, and environmental context^7^. Recent studies show that such agents can reproduce human-like patterns in controlled social science experiments and surveys^8,9^, underscoring their potential as a data-driven framework for behavioral simulation. However, it remains unclear whether LLM-based agents can replicate policy adherence under complex real-world conditions, where behavioral responses are shaped jointly by dynamic disease risks, varying levels of policy interventions, and potential biases inherited from pretraining data. Here, we test whether LLM agents can reproduce observed mobility patterns in real-world foot-traffic data during the early stage of the COVID-19 pandemic, reproduce heterogeneity across cities and place categories, and potentially serve as a generative behavioral layer for epidemic modeling.

## Results

We conducted simulation experiments in three U.S. cities (Fig. 1a), including Boston, Denver, and San Antonio, selected to capture diverse geographic, economic, and cultural contexts (Appendix Section 1.3). For each city, we instantiated 300 LLM agents whose demographic profiles matched city-specific marginal distributions of age, race, income, and occupation. To approximate the information environment of spring 2020 without explicitly naming the historical period or pathogen identity, agents were prompted with information about an unidentified emerging respiratory virus scenario and weekly updates on national and local disease risks, together with policy measures implemented in that city at the corresponding time points. For each POI category, agents generated the expected percentage change in visitation relative to their baseline behavior, defined as their normal activity level in the absence of a pandemic. By embedding policy scenarios within the prompts, we elicited mobility decisions conditioned on the evolving outbreak situation, individual characteristics, and broader socioeconomic and cultural contexts. In this study, we evaluated all policies implemented in each city according to their observed temporal sequence, focusing on 3 categories of POIs (e.g., restaurants and bars, retail stores, and art and entertainment venues). To assess variability across models, we conducted experiments with three LLMs: GPT-4.1, Gemini-2.5-Pro, and Grok-4.1-Fast.

**Figure 1.**
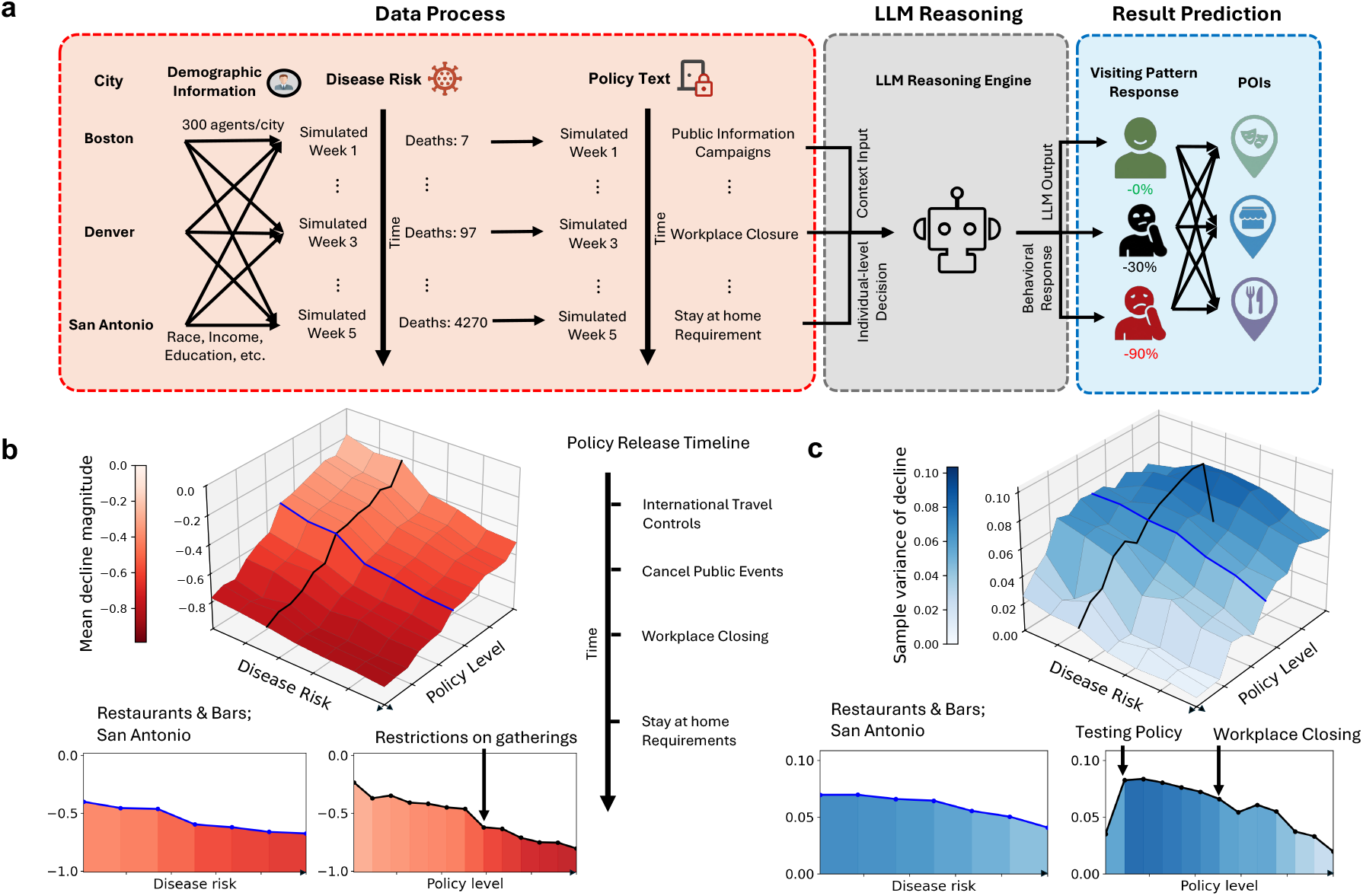
Overview and sensitivity analysis of the LLM-based mobility simulation framework. **a**, Framework illustration. The overall pipeline integrates demographic context and situational inputs (e.g., disease risk and policy interventions) into a contextual reasoning engine, where an LLM generates agent-level behavioral responses, producing visiting pattern adjustments across POI categories. **b**, Mean response surface for Restaurants and Bars in San Antonio. The 3D heatmap shows the joint average mobility response of simulated agents to varying disease risk and policy levels. The corresponding 2D slices show the marginal patterns, illustrating how average visitation changes with policy levels when disease risk is held fixed, and how it changes with disease risk when policy level is held fixed. The vertical timeline shows the temporal order of major policies enacted in San Antonio, reflecting the increasing policy stringency tested in the simulations. **c**, Variance response surface for Restaurants and Bars in San Antonio. A 3D heatmap shows the joint variation in agent-level mobility responses across disease-risk and policy conditions. The corresponding 2D slices show the marginal variance patterns with one input varied while the other is held fixed, illustrating how behavioral heterogeneity changes across the policy–risk space. Equivalent results for the heatmap of POI types Arts & Entertainment and Retail are shown in Appendix Supplementary Figs. 1 and 2.

We first simulated mobility responses across combinations of infection risk and policy stringency. The simulated responses were sensitive to both epidemic severity and policy intensity within the range observed in March 2020. For restaurant and bar visits, mean mobility declined monotonically with increasing disease risk and policy stringency (Fig. 1b). Marginal response curves further indicate that, within this empirical range, policy intensity exerts a stronger effect on mobility reduction than disease risk (Fig. 1b). Among all interventions, workplace closures and stay-at-home orders produced the largest declines, consistent with evidence that these measures are the primary drivers of population-level behavioral change during acute outbreak phases^10^. Behavioral heterogeneity among agents was low in the absence of interventions and disease risk, increased with early policy introduction under low disease burden, and declined as policy stringency or disease risk intensified (Fig. 1c). This pattern suggests that agent-level differences are most pronounced under moderate decision pressure, when responses depend on how agents interpret weaker policy signals through their socioeconomic profiles. By contrast, stringent mandates or high disease burden impose strong constraints, leading to more uniform reductions in mobility.

Focusing on real-world disease situations and policies in spring 2020, the average LLM predictions approximated observed median city-level mobility reductions across POI categories (Fig. 2a). Table 1 summarizes model performance across cities, POI categories, and LLM architectures, using the Jensen–Shannon divergence and median differences between simulated and observed mobility changes. Detailed formulas for these metrics are defined in Appendix Section 2.1. Individual LLM exhibited distinct biases. Gemini tended to produce overly adherent responses following policy announcements, although it performed most accurately during the early phase of the pandemic (Appendix Supplementary Figs. 6-16). GPT exhibited an intermediate level of conservatism between Gemini and Grok, whereas Grok produced the least conservative responses, generating the smallest reductions in simulated mobility. The ensemble achieved the best average rank across cities, suggesting that ensembling can reduce dependence on any single model’s behavioral prior and improve the stability of population-level estimates. Performance was strongest for Retail, whereas Arts & Entertainment showed substantial discrepancy, with simulated responses generally overestimating mobility reductions. We further examined responses following the specific policy “gatherings of more than 10 people prohibited.” LLM agents correctly reproduced the relative ordering of mobility reductions across place categories in all three cities (Fig. 2b–d), indicating sensitivity to context-specific policy effects. These results show that LLM agents can approximate city-level median mobility responses without empirical fine-tuning, particularly for routine activities, while remaining less reliable for discretionary behaviors shaped by subjective preferences^11^.

**Table 1.** Comparison of LLM-agent performance across cities and POI categories. Jensen-Shannon divergence measures distributional agreement between simulated and observed mobility-change distributions, while median difference measures the absolute difference between simulated and observed median responses. Lower values indicate better performance. Rankings within each city–POI–metric combination are shown in parentheses, and the best performing value is bolded.

| City | Model | Restaurants & Bars |  | Retail |  | Arts & Entertainment |  | Rank |
| --- | --- | --- | --- | --- | --- | --- | --- | --- |
|  |  | JS Div. ↓ | Median Diff. ↓ | JS Div. ↓ | Median Diff. ↓ | JS Div. ↓ | Median Diff. ↓ |  |
| Boston | GPT-4.1 | 0.225 (3) | 0.085 (2) | 0.238 (3) | 0.141 (3) | 0.374 (3) | 0.292 (2) | 2.667 |
|  | Gemini-2.5-Pro | 0.241 (4) | 0.278 (4) | 0.147 (2) | **0.043** (1) | 0.428 (4) | 0.484 (4) | 3.167 |
|  | Grok-4.1-Fast | 0.198 (2) | 0.104 (3) | 0.277 (4) | 0.271 (4) | **0.274** (1) | **0.206** (1) | 2.500 |
|  | Ensemble | **0.165** (1) | **0.079** (1) | **0.143** (1) | 0.133 (2) | 0.301 (2) | 0.304 (3) | **1.667** |
| Denver | GPT-4.1 | 0.257 (3) | **0.107** (1) | 0.235 (3) | 0.063 (2) | 0.413 (3) | 0.306 (2) | 2.333 |
|  | Gemini-2.5-Pro | 0.262 (4) | 0.265 (4) | 0.158 (2) | 0.132 (3) | 0.457 (4) | 0.486 (4) | 3.500 |
|  | Grok-4.1-Fast | 0.218 (2) | 0.119 (3) | 0.251 (4) | 0.182 (4) | **0.319** (1) | **0.224** (1) | 2.500 |
|  | Ensemble | **0.178** (1) | 0.115 (2) | **0.133** (1) | **0.061** (1) | 0.345 (2) | 0.326 (3) | **1.667** |
| San Antonio | GPT-4.1 | 0.174 (3) | **0.098** (1) | 0.104 (2) | **0.051** (1) | 0.284 (3) | 0.159 (2) | **2.000** |
|  | Gemini-2.5-Pro | 0.209 (4) | 0.198 (4) | 0.142 (4) | 0.166 (4) | 0.385 (4) | 0.341 (4) | 4.000 |
|  | Grok-4.1-Fast | 0.137 (2) | 0.102 (2) | 0.113 (3) | 0.092 (3) | **0.196** (1) | **0.129** (1) | **2.000** |
|  | Ensemble | **0.106** (1) | 0.108 (3) | **0.066** (1) | 0.067 (2) | 0.248 (2) | 0.185 (3) | **2.000** |

**Figure 2.**
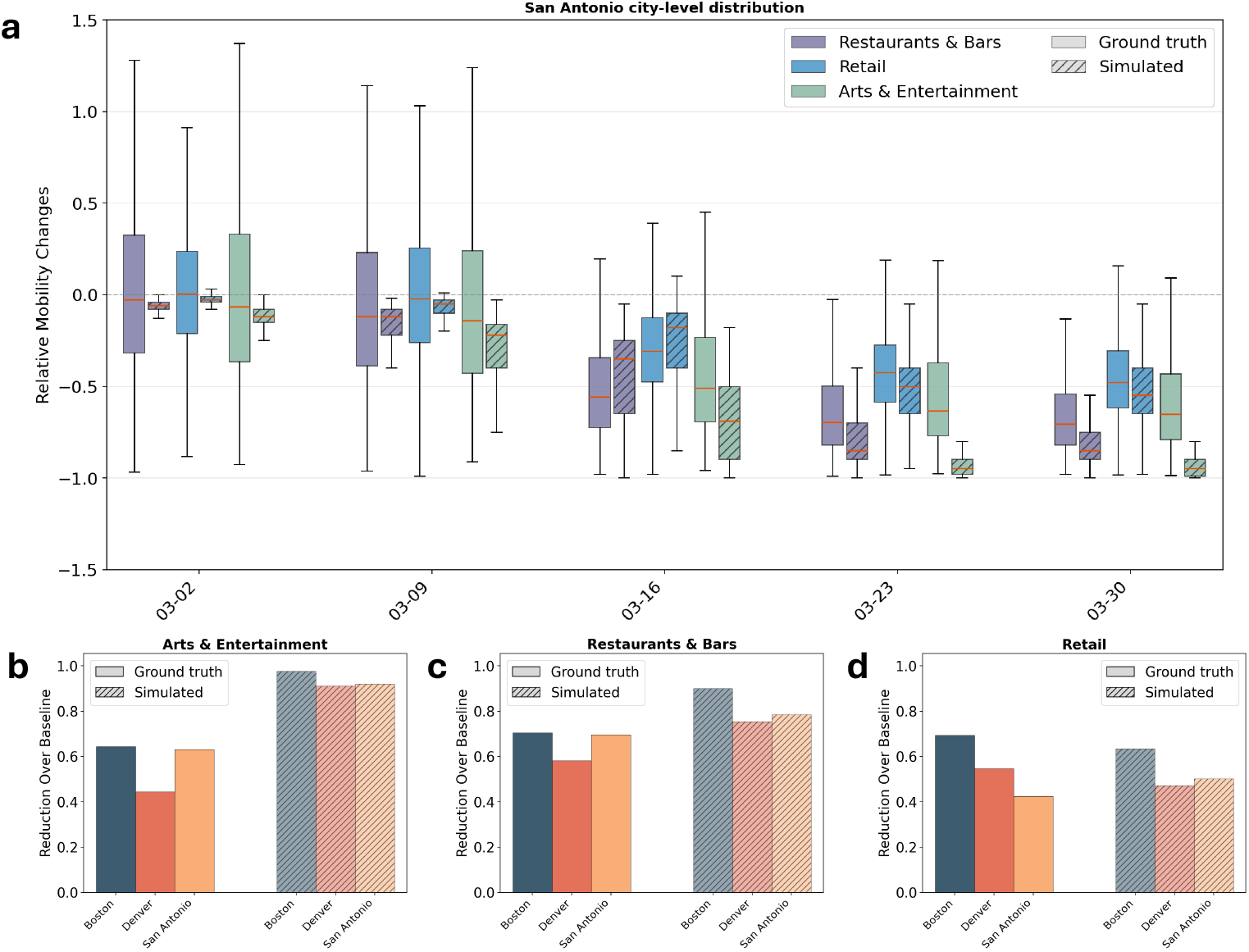
City-level comparison between observed and simulated mobility responses across POI categories. **a**, Distributional comparison for San Antonio. The box plot shows the city-level distribution of visitation changes in San Antonio across five dates, comparing observed and simulated outputs for all three POI categories. Solid box plots represent the observed real-world reduction distributions across CBGs, and dashed box plots represent the simulated reduction distributions among agents. Boxes represent the interquartiles, and whiskers show the 95% CIs. Equivalent results for Boston and Denver, including results for different models, are presented in Appendix Supplementary Fig. 6-16. **b–d**, Visitation reduction relative to the baseline across Boston, Denver, and San Antonio for the same three POI categories: **b**, Arts & Entertainment, **c**, Restaurants & Bars, and **d**, Retail. Bars compare observed mobility reduction across the three cities on the week immediately following the policy “Gatherings of more than 10 people prohibited” was enacted in each city. Bar chart results for other policy examples are shown in Appendix Supplementary Figs. 17 and 18.

Although LLM agents accurately reproduced city-level median mobility reductions, they systematically underestimated the variation in mobility observed across census block groups (CBGs), reflecting a tendency toward over-compliance with population norms (Fig. 2a). Simulated distributions of mobility changes were narrower than the observed ranges, particularly in the early weeks when real-world behavioral responses were highly heterogeneous. Increasing the number of agents did not resolve this mismatch (Appendix Supplementary Fig. 3-5). This pattern is consistent with the response-variance analysis in Fig. 1c. When external constraints were weak, LLM agents tended to generate responses close to the population average rather than the broader range of behaviors observed across communities. The simulated distribution also tended to show steeper declines than the observed distribution, indicating that LLM agents were more policy-adherent than real populations. This over-compliance is consistent with known limitations of LLM-based behavioral simulation, including social desirability^12^ and normative response biases^13^, in which models lean toward what an idealized person should do rather than what heterogeneous populations actually do. This finding suggests that current LLMs approximate average compliance more reliably than they simulate the heterogeneous behavioral responses that shape neighborhood-scale variation.

## Discussion

Our results demonstrate that LLM agents can reproduce city-level average adherence to NPIs across multiple cities and the three examined POI categories. For epidemic models that rely on aggregated, population-level mobility patterns^6^, LLM agents can be integrated as a behavioral layer to more realistically represent the effects of NPIs. However, the framework underestimates within-city variation in mobility, indicating the need for further calibration or fine-tuning to better represent uncertainty and subgroup-level heterogeneity. We also identify systematic biases across LLMs, which may reflect model-specific behavioral priors shaped by training data and alignment procedures. Although multi-model ensembling mitigates these effects, it does not eliminate them. These findings underscore the importance of rigorously validating and aligning LLMs with demographically and culturally diverse populations^14^ to support the equitable deployment of AI-based modeling approaches.

A number of limitations should be noted. First, due to data availability, we focused on three place categories with relatively rich foot-traffic records. Performance in other settings, such as workplaces and schools, warrants further investigation. Second, foot-traffic data may be subject to sampling biases across subpopulations. Our use of relative mobility changes partially mitigates this issue. Third, we focus on mobility responses during the initial phase of a pandemic. Future work should examine whether LLM agents can capture non-stationary behavioral dynamics, including reopening, pandemic fatigue^15^, evolving risk perception, and sequential decision-making with shifting information. Because contemporary LLMs may contain extensive information about COVID-19 in their training data, future work should also test whether similar zero-shot behavior emerges in less historically recognizable or prospective outbreak scenarios. Despite these limitations, LLM-based behavioral simulation offers a practical alternative to fixed scenario-based assumptions in epidemiological models^16^, enabling context-sensitive representations of human behavior. Coupled with mobility-driven epidemic models, this framework may support more responsive forecasting and intervention design tailored to specific socioeconomic and cultural contexts.

## Methods

In this work, we aim to simulate changes in human visiting patterns in response to evolving public health policies and COVID-19 severity. Simulations were conducted at the agent level, where agents were instantiated from multiple cities, and for each city, the demographic composition of agents was constructed to match its marginal population distributions. Each agent’s behavior is modeled by integrating individual demographic attributes with temporally varying pandemic conditions, characterized by confirmed case counts and death statistics at the state and national levels, as well as officially published policy interventions in the corresponding state up to the simulated date.

Conditioned on these factors, large language models were employed to infer the degree of visitation change relative to pre-pandemic baselines. Predictions are generated for five selected weeks, each representing a different stage of early-pandemic progression, and results are aggregated across multiple LLM backbones to mitigate model-specific biases. The framework ultimately produces agent-level visitation adjustments across diverse POI categories over time, which are then aggregated to recover city-level mobility patterns and their evolution across different urban environments.

### Data

Our study incorporated four categories of data: 1) demographic and socioeconomic data for constructing representative agent profiles, 2) human mobility data as empirical benchmarks, 3) epidemiological data describing pandemic severity, and 4) policy records describing officially reported interventions in effect up to each simulated date. Detailed data processing methods are reported in Appendix Section 1.

#### Agent demographic background

We utilize census block group–level demographic and socioeconomic data obtained from the U.S. Census Bureau’s American Community Survey (ACS)^17^, encompassing attributes including race, gender, age, household income, education level, and occupation. All demographic inputs in this study are based on the 2020 five-year ACS estimates.

#### Human mobility data

To establish real-world mobility patterns as a reference benchmark, we adopt the Advan Weekly Patterns Plus dataset^18^. Our analysis considers three U.S. metropolitan areas chosen to capture variation in urban scale and spatial configuration: Boston, a dense and transit-oriented city with a compact structure; Denver, a mid-sized metropolitan area with a more expansive and decentralized layout; and San Antonio, a large southern city characterized by relatively lower density and distinct urban form. The dataset contains aggregated, anonymized location signals collected from millions of mobile devices and provides weekly visit flow summaries from census block groups (CBGs) to different categories of points of interest (POIs). POI aggregation rules are provided in the Appendix Section Supplementary Table 1.

We construct the mobility benchmark to evaluate our framework’s ability to capture mobility changes under evolving pandemic conditions. Starting from weekly origin–destination (OD) flows, we measured visits from each home CBG to individual POIs within each city and then aggregated POIs into the three study categories: 1) Restaurants and Bars, 2) Retail, and 3) Arts and Entertainment. For each city and POI category, we define baseline mobility as the average weekly visitation level across four pre-pandemic weeks between January and February 2020. We then compare these baseline visitation levels with weekly observations in March 2020 to compute relative mobility changes, separately for each city–POI pair. These relative changes served as the empirical benchmark for evaluating simulated visitation adjustments. The mobility records were used only for evaluation and were not provided to agents or used for model calibration.

#### Disease severity information

We utilize epidemiological data from the JHU CSSE COVID-19 dashboard^19^, which provides records of cumulative confirmed COVID-19 cases and deaths over time at both the state and national levels. We use these data to characterize pandemic severity at each simulated date; a full detailed disease timeline is shown in Appendix Section Supplementary Table 2.

#### Policy data

To characterize the policy environment, we draw upon pandemic-related policy records from the Oxford COVID-19 Government Response Tracker^20^, which systematically records government interventions and their temporal deployment. This resource captures a range of policy actions, including stay-at-home mandates, guidance on social distancing practices, and targeted restrictions across sectors such as education, transportation, and non-essential services. For each simulated week, we summarized the policy measures in effect as standardized text descriptions and included these descriptions in the agent prompts. A detailed policy timeline is shown in Appendix Section Supplementary Table 3.

### Simulating policy response

In this section, we outline a simulation pipeline consisting of agent sampling, prompt construction, model querying, output collection, and evaluation. Below, we describe each component of the pipeline and the metrics used to compare simulated mobility responses with observed mobility changes.

#### LLM Agent Setup

To capture demographic heterogeneity at the individual level, we employed a hybrid LLM-IPF framework to construct representative generative agents. Building upon traditional Iterative Proportional Fitting (IPF)^21^, we incorporate LLM-derived auxiliary constraints to supplement census-based marginal distributions and improve the plausibility of joint agent attributes. For each city, agents were initialized with a persona profile consisting of demographic and socioeconomic attributes (e.g., sex, age, race, and neighborhood-level income and education). The joint distribution of these attributes was estimated using city-specific census marginals and auxiliary constraints. For agent instantiation, we generated 300 agents per city across three representative urban areas: Boston, Denver, and San Antonio, to balance computational efficiency and behavioral diversity. Details of the agent sampling procedure are provided in Appendix Section 2.2.

#### Prompt Setup

To simulate realistic agent-level mobility responses under pandemic conditions, we constructed a structured prompting framework that integrates individual, policy, and epidemiological context. The prompt was composed of three primary components. First, the demographic context defined the agent’s persona, including attributes such as age, occupation, and socioeconomic background. Second, the policy timeline encoded the sequence, timing, and intensity of public health interventions. Third, the disease situation summarized the current epidemiological conditions, including case trends and associated risks. Beyond these core components, we incorporated auxiliary sections to improve reasoning fidelity and task clarity. In particular, a helper module explicitly specified the prediction objective, ensuring consistency in expected outputs, while an additional chain-of-thought guidance module encouraged step-by-step reasoning grounded in the provided context. A detailed prompt example is provided in Appendix Section 2.3.

#### Prompt Sensitivity Analysis

To assess the robustness of our ensemble model to prompt formulation, we conducted a prompt sensitivity analysis using five distinct rephrased variants of the original prompt. Each variant preserved the core epidemiological intent while varying phrasing, structure, and terminology. For each prompt variant, we ran the ensemble model pipeline using Denver as the experimental setting, generating complete output distributions across all simulation runs. The resulting distributions were then compared across variants to evaluate consistency and stability. Our analysis showed that the ensemble model produced largely stable predictions across prompt reformulations, suggesting limited sensitivity to minor linguistic variations in the input prompt. Detailed prompt sensitivity analyses are shown in Appendix Supplementary Fig. 19-21.

#### Model Selection

We evaluated three distinct LLMs: GPT-4.1, Gemini-2.5-Pro, and Grok-4.1-Fast to model visiting pattern changes conditioned on agent demographics, evolving disease conditions, and contemporaneous policy interventions on simulated dates. Each model was queried using the same agent profiles, policy context, disease-severity information, and output format to ensure comparability across model backbones. To reduce dependence on any single model backbone, we adopted an ensemble strategy that combines outputs from all three models, using the aggregated predicted visitation change across models as the ensemble estimate. Model-specific and ensemble outputs were retained separately for subsequent evaluation against observed mobility changes.

#### Evaluation of Simulation Results

We evaluated the simulation results by comparing the observed and simulated distributions of relative visitation changes for each city, week, and POI category. The observed mobility data were available only at the CBG level. In contrast, the simulation generated agent-level visitation changes. We therefore compared the two distributions under the assumption that the simulated agent population provides an individual-level approximation to the range of behavioral responses that, when aggregated in the real data, appears as variation across CBGs. This comparison is not intended to establish a one-to-one correspondence between agents and CBGs, but rather to assess whether the simulated responses recover the central tendency and dispersion of observed mobility changes within the same city–week–POI category setting. To compare the observed and simulated distributions, we used Jensen–Shannon (JS) divergence as a symmetric measure of distributional similarity. In addition, we computed the median difference between the two distributions to capture deviations in central tendency. Together, these two metrics capture complementary aspects of model performance: JS divergence evaluates distributional agreement, whereas median difference evaluates the accuracy of the central response. Both metrics were computed separately for each city, POI category, simulated date, and model configuration. Formulas for both metrics are provided in Appendix Section 2.1.

## Data availability

Demographic data are publicly available from the U.S. Census Bureau American Community Survey (https://www.census.gov/data.html). COVID-19 case and death data are publicly available from the Johns Hopkins University Center for Systems Science and Engineering COVID-19 dashboard (https://github.com/cssegisanddata/covid-19), and policy data are publicly available from the Oxford COVID-19 Government Response Tracker (https://github.com/OxCGRT/covid-policy-tracker). The mo- bility data were derived from Advan Research Weekly Patterns Plus data accessed through the University of Virginia’s institutional license via Dewey. Raw Advan data cannot be publicly redistributed. Researchers seeking access to the original mobility data should obtain it directly from Dewey under the applicable licensing terms (https://www.deweydata.io/).

## Code availability

All code used to implement the models can be obtained from: GitHub repository (Policy Agent).

## Acknowledgements

This work was supported by NSF Award ID 2229605 (S.P.) and National Institute of General Medical Sciences R35GM156799 (S.P.). Its contents are solely the responsibility of the authors and do not necessarily represent the official views of the funding agencies.

## Competing interests

The authors declare no competing interests.

## Notes

### Competing Interest Statement

The authors have declared no competing interest.

### Author Declarations

All datasets used in this study, including epidemiological data from the Johns Hopkins University CSSE dashboard, policy data from the Oxford COVID-19 Government Response Tracker, and mobility data, were aggregated at the population level. No individual-level data were used in this research.

