## Supplementary Information for "Simulating population compliance with pandemic interventions using large language models"

**Runzhou Liu[1], Claire Jong[2], Haoyang Li[1], Yiming Cao[3], Qing Yao[3], Teresa Yamana[4], Sen Pei[3,*], and Hongru Du[1,*]**

[1]Department of Systems & Information Engineering, University of Virginia, Charlottesville, VA, USA
[2]Department of Computer Science, Columbia University, New York, NY, USA
[3]Department of Environmental Health Sciences, Mailman School of Public Health, Columbia University, New York, NY, USA
[4]Columbia Climate School, Columbia University, New York, NY, USA
[*] (SP), (HD)

**References** **17**

### 1 Supplementary Data

#### 1.1 Classification and Aggregation of POI

We organized POIs defined by the North American Industry Classification System (NAICS)[1] into three functional categories to better capture residents' primary activity spaces. This grouping was based on aligning economic sectors that share similar mobility patterns and underlying travel purposes. For example, sectors such as Full-Service Restaurants and Drinking Places were consolidated into a single category, "Restaurants and Bars". By abstracting detailed industrial classifications into broader functional groups, this approach simplifies the semantic structure of the environment, enabling generative agents to reason more effectively about essential daily activities rather than fragmented sector-level distinctions. The complete mapping is provided in the Supplementary Table1.

**Supplementary Table 1: Mapping of NAICS sector codes to selected functional activity categories.** The table groups POIs into three core categories based on NAICS sector prefixes, capturing key urban activity spaces relevant to mobility behavior.

| NAICS Sector Code | NAICS Sector Full Name | Aggregated Category | Logic of Aggregation |
|---|---|---|---|
| 44, 45 | Retail Trade | Retail | Commercial logistics and consumer purchasing centers. |
| 7224, 722511 | Drinking Places, Full-Service Restaurants | Restaurants & Bars | Social dining, food consumption, and nightlife-related activities. |
| 7111, 71211, 7132, 71394, 71395 | Performing Arts Companies, Museums, Gambling Industries, Fitness and Recreational Sports Centers, Bowling Centers | Arts & Entertainment | Leisure, cultural engagement, and recreational activities. |

#### 1.2 Data Process

Using the U.S. Census Bureau's American Community Survey (ACS)[2], the data processing pipeline constructs structured demographic marginals at the census block group (CBG) level by aggregating raw census attributes into semantically meaningful categories. For race, population counts are directly grouped into three categories: White, Black, and Other Race. For education, we restrict to working-age adults and aggregate attainment into five levels: High school and lower, Associate's, Bachelor's, Master's, Professional School, and Doctorate degrees. For household income, values are consolidated into three tiers: Low, Medium, and High, reflecting standard U.S. income brackets. For age, gender-specific census marginals are first combined within each gender and then summed into seven working-age brackets: 21–29, 30–39, 40–49, 50–59, 60–69, 70–79, and 80+. For occupation, the original 23 detailed categories are compressed into seven broader functional groups to reduce sparsity and improve interpretability. The final occupation categories include:

- High-Skill Professional & Technical
- Education & Public / Social Services
- Healthcare
- Creative & Cultural
- Service Occupations
- Clerical & Sales
- Blue-Collar / Manual Labor

Each CBG row represents a small geographic unit. City-level agent generation requires combining all CBGs within a city boundary into a single representative demographic profile for each city.

#### 1.3 Study Site Selection

To enable meaningful comparison of LLM-assessed policy adherence across locations with varying political contexts, we selected three metropolitan areas: Boston, Massachusetts; Denver, Colorado; and San Antonio, Texas. We used these locations to represent a spectrum of political orientations while maintaining comparability across key epidemiological and policy dimensions. Candidate cities were initially drawn from a geographically and politically diverse pool of ten U.S. metropolitan areas, chosen to span regional variation and a range of political leanings as implemented by the conservatism score developed by Tausanovitch and Warshaw[3]. Boston was classified as liberal, Denver as moderate, and San Antonio as conservative based on this metric. Epidemiological comparability was assessed using county-level cumulative confirmed COVID-19 case and death data from the Johns Hopkins University Center for Systems Science and Engineering (CSSE) repository[4], with per-capita figures used to identify locations with broadly similar outbreak severity during the study period. Policy timing was characterized using the Oxford COVID-19 Government Response Tracker[5]. Key policy actions examined included stay-at-home orders, closures of non-essential businesses, and restaurant restrictions. State- and county-level mobility data reported by Badr et al.[6] were additionally consulted to assess whether the selected locations exhibited meaningful variation in population movement during the study period. The final cities were selected from among the candidate locations on the basis of having the most similar stay-at-home order issue dates and durations, facilitating temporal alignment in the policy adherence analysis while still representing meaningfully distinct political contexts.

#### 1.4 Pandemic Timeline

We utilize epidemiological data from the JHU CSSE COVID-19 dashboard[4]. The pandemic timeline captures the rapid escalation of COVID-19 confirmed cases and deaths across the United States from late February through the end of March 2020. In the early weeks, case counts at both the state and national levels remained extremely low, with most states reporting single digit infections. However, by mid-March, following the WHO's declaration of a global pandemic on March 11 and the U.S. national emergency on March 13, case trajectories steepened sharply. By March 30, Massachusetts had recorded 8,647 cases and 47 deaths, Colorado 2,467 cases and 50 deaths, Texas 3,173 cases and 65 deaths, and the nation as a whole had surpassed 165,000 confirmed infections and over 4,200 deaths. Detailed statistical results for different cities on different dates are shown in Table 2.

**Supplementary Table 2:** COVID-19 confirmed cases and deaths across states and the United States over time.

| City / Region | 2/24/2020 | | 3/9/2020 | | 3/16/2020 | | 3/23/2020 | | 3/30/2020 | |
|---|---|---|---|---|---|---|---|---|---|---|
| | Conf. | Deaths | Conf. | Deaths | Conf. | Deaths | Conf. | Deaths | Conf. | Deaths |
| Boston / MA | 2 | 0 | 96 | 0 | 512 | 0 | 2,899 | 2 | 8,647 | 47 |
| Denver / CO | 0 | 0 | 12 | 0 | 161 | 2 | 718 | 7 | 2,467 | 50 |
| San Antonio / TX | 0 | 0 | 13 | 0 | 85 | 0 | 764 | 11 | 3,173 | 65 |
| United States | 17 | 1 | 630 | 24 | 4,638 | 97 | 45,952 | 786 | 165,282 | 4,270 |

#### 1.5 Policy Timeline

We take use of the Oxford COVID-19 Government Response Tracker dataset[5]. The policy timeline documents the sequence of policy interventions enacted across Boston (Massachusetts), Denver (Colorado), and San Antonio (Texas) between late February and March 30, 2020. Policies progressed in a consistent pattern across all three cities: public information campaigns and targeted testing guidelines came first in late February, followed by event cancellations and nursing home restrictions in early-to-mid March. After the twin news shocks of the WHO pandemic declaration and the U.S. national emergency[7], each state rapidly enacted school closures, restaurant and bar bans, gathering limits, and ultimately stay-at-home orders. A detailed policy timeline is shown in Table 3.

**Supplementary Table 3:** Policy timeline across cities during early COVID-19 response.

| Policy / Event | Boston (MA) | Denver (CO) | San Antonio (TX) |
|---|---|---|---|
| Nursing home & long-term care facility (LTCF) visitor restrictions and hygiene protocols recommended | 2/26/2020 | — | — |
| Public information campaigns | 3/4/2020 | 2/27/2020 | — |
| Testing policy to patients meeting specific clinical and exposure criteria | 2/28/2020 | 3/2/2020 | 3/4/2020 |
| Contact tracing initiated for confirmed COVID-19 cases | — | 3/5/2020 | 3/4/2020 |
| Cancellation of all public events and gatherings required | 3/13/2020 | 3/19/2020 | 3/11/2020 |
| Gatherings of more than 1,000 people prohibited | 3/13/2020 | 3/12/2020 | 3/13/2020 |
| Closure of schools required in affected areas or for select grade levels | 3/12/2020 | 3/16/2020 | 3/16/2020 |
| Workplace closure for certain sectors | — | 3/15/2020 | 3/16/2020 |
| Gatherings of more than 100 people prohibited | 3/17/2020 | 3/12/2020 | — |
| Protection of elderly people in LTCF | 3/12/2020 | 3/12/2020 | 3/21/2020 |
| Coordinated, state-wide public information campaign on COVID-19 prevention launched | 3/13/2020 | 3/16/2020 | 3/27/2020 |
| Gatherings of more than 10 people prohibited | 3/24/2020 | 3/20/2020 | 3/17/2020 |
| Required closing of all schools and educational institutions at all levels, statewide | 3/23/2020 | 3/30/2020 | 3/20/2020 |
| Stay at home advisory issued; residents strongly urged to remain home for non-essential activities | 3/23/2020 | — | 3/20/2020 |
| Stay at home requirement, except essential trips | — | 3/24/2020 | 3/23/2020 |
| All non-essential businesses and workplaces required to close | 3/24/2020 | 3/24/2020 | — |
| Reduction or suspension of public transportation services | 3/27/2020 | 3/29/2020 | — |
| Travel between regions or cities discouraged; residents advised not to cross regional boundaries | 3/27/2020 | — | — |
| Restrictions on internal movement | — | — | 3/28/2020 |
| NEWS: WHO Declares COVID-19 a Global Pandemic | 3/11/2020 | 3/11/2020 | 3/11/2020 |
| NEWS: U.S. National Emergency Declared + European Travel Ban | 3/13/2020 | 3/13/2020 | 3/13/2020 |

#### 2 Supplementary Methods

##### 2.1 Evaluation Metrics

We evaluate simulation performance using two complementary metrics that capture both distributional similarity and central tendency differences. Jensen–Shannon (JS) divergence is adopted as a symmetric and robust measure for comparing distributions. The equation for JS divergence can be seen in Eq. 1

$$JS(P \parallel Q) = \frac{1}{2} KL(P \parallel M) + \frac{1}{2} KL(Q \parallel M), \quad \text{where } M = \frac{1}{2}(P+Q) \tag{1}$$

The absolute median difference quantifies deviation in typical values between simulated and ground-truth distributions. The equation for median difference can be seen in Eq. 2

$$|\text{median}(P) - \text{median}(Q)| \tag{2}$$

##### 2.2 Sampling Method

Standard iterative proportional fitting (IPF)[8] operates only on 1-D marginals. Real populations, however, exhibit strong inter-variable correlations. For example, education level correlates with income. Therefore, we introduce a two phase LLM+IPF method that augments IPF with an LLM that infers these dependencies and steers generation to honor both marginal and joint constraints.

In Phase 1 variable dependency inference, the LLM is prompted with observed marginals and available joint distributions (gender × age and gender × occupation) to infer additional high impact variable dependencies. The model identifies the most relevant variable pairs and estimates approximate joint distributions for these pairs. To ensure robustness, the inference process is repeated multiple times, and the outputs are aggregated through ensemble averaging, producing stable dependency subsets and smoothed joint estimates. These inferred joints complement the real census joints and are treated as soft constraints with reduced weight, reflecting their uncertainty.

In Phase 2 iterative constrained generation, agents are generated through an iterative loop guided by both marginal and joint constraints. At each step, the system computes discrepancies between current and target distributions, and the LLM proposes batches of agents that help reduce these gaps by specifying partial attribute assignments. The remaining attributes are filled via residual-weighted sampling to prioritize underrepresented categories. This process continues until the convergence criteria are met. A final correction phase further refines the population by removing overrepresented agents and regenerating replacements, ensuring that the synthesized population closely matches both marginal distributions and key dependency structures.

##### 2.3 Prompt Template

The prompt is constructed by demographic information, situation information, and task-guided helper information listed below:

**Background Information**

You are a resident living in **<City Name>**, **<State>**. *City State Introduction*
**[Demographic Context]**
You are a **<Age>**, **<Race>**, **<Gender>**, with a **<Education>** degree, living in a **<Household Income>** household. Your occupation is in **<Occupation>**. The total population of your city is **<Total Population>**.
**[Policy Timeline]**
*Policy Timeline*
**[Disease Situation]**
A new infectious disease, temporarily designated as "Novus-Pathogen", is currently active. It spreads via respiratory droplets and surface contact, with symptoms similar to influenza (fever, cough, fatigue). Severe cases may lead to pneumonia or death, especially for vulnerable populations.
*Disease Situation Stats*

**Your Task: Predict Mobility Changes**

Based on your demographic profile, policy environment, and disease situation, predict how your visitation behavior will change compared to normal for three POI categories:

- Restaurants & Bars
- Retail
- Arts & Entertainment

**Reasoning Process**

Before making your prediction, you must reason step-by-step:

1. **Policy compliance reasoning:** Consider your personal attributes (age, race, gender, education, income). Decide whether you would comply with the policies in the timeline and how they influence your mobility behavior.
2. **Disease awareness:** If policies are weak or absent, consider infection risk using confirmed cases and death counts. Low risk should not overly influence behavior, but should still be considered.
3. **Mobility constraint:** Stay-at-home advisories are not strict lockdowns; essential trips (e.g., groceries, medicine) are still allowed.
4. **Final decision:** Based on the above reasoning, determine your visitation change for each POI category.

The output should contain two sections for each of the poi types:

**Reasoning**: your chain of thought as described in Reasoning Process, including policy compliance reasoning and final decision.

**Number output**: A number indicate your percentage change of visitation.

If the visitation tend to increase around 15% compare to normal, the percentage number should be positive. (Ex: 0.15)
If the visitation tend to decrease around 16% compare to normal, the percentage number should be negative. (Ex: -0.16)

Return your answer in the following JSON format:

```
{
   "Restaurants_Bars_reasoning": "...",
   "Restaurants_Bars_change": <float>,

   "Retail_reasoning": "...",
   "Retail_change": <float>,

   "Arts_Entertainment_reasoning": "...",
   "Arts_Entertainment_change": <float>,
}
```

### 3 Supplementary Results

#### 3.1 Heat Map for all POIs

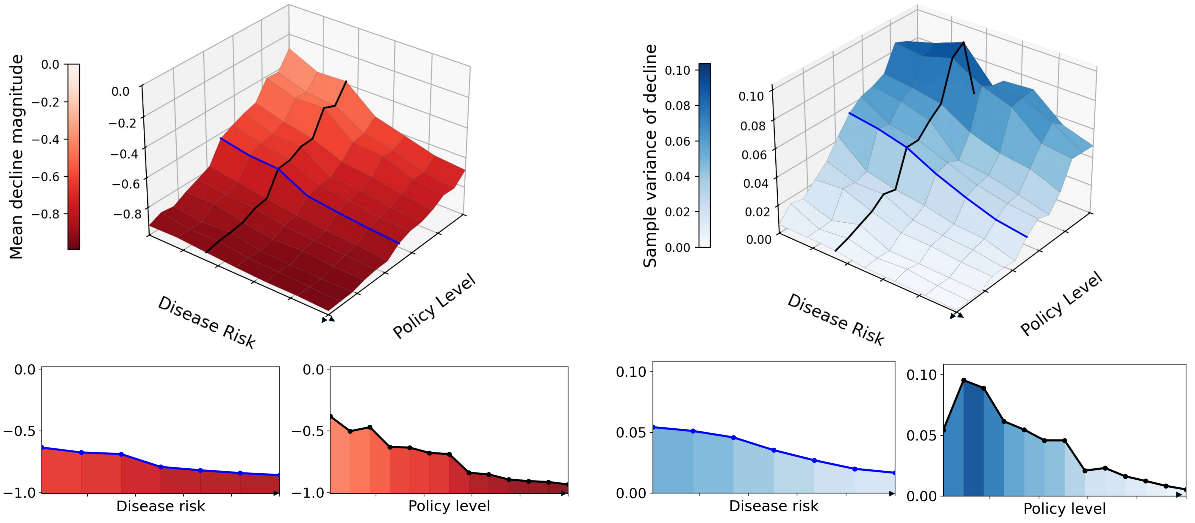

**Supplementary Figure 1: Sensitivity analysis of simulated mobility responses for Arts & Entertainment. Left**, mean response. A 3D heatmap shows the average visitation reduction, with a clear monotonic decline as disease risk increases and policies become more restrictive. The accompanying 2D slices highlight marginal effects along each dimension. **Right**, variance response. A 3D heatmap depicts the sample variance of visitation changes, capturing the evolution of behavioral heterogeneity. The 2D projections indicate that variance decreases under higher risk and stricter policies, suggesting more uniformly reduced mobility across agents.

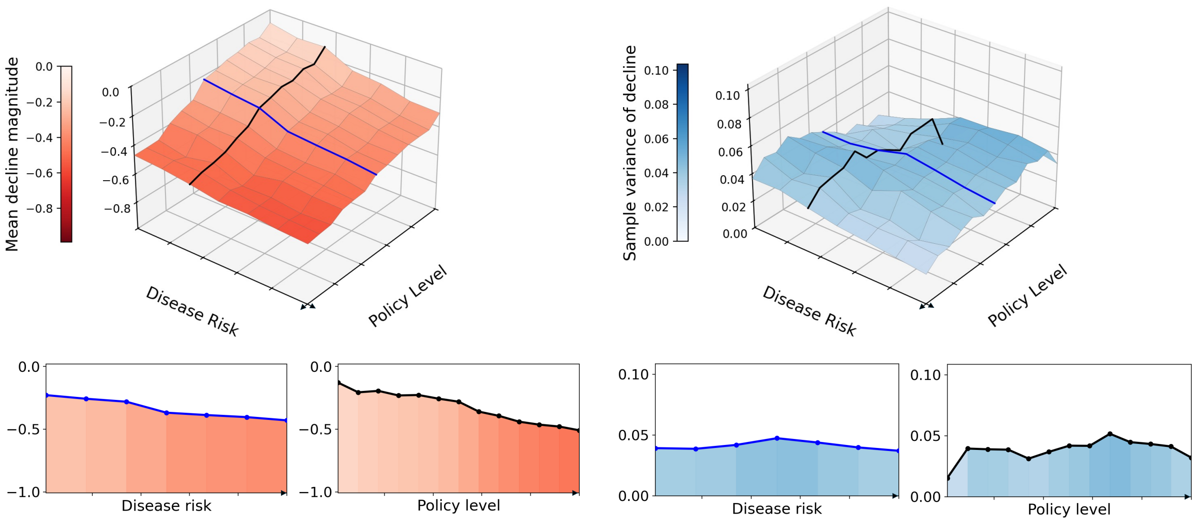

**Supplementary Figure 2: Sensitivity analysis of simulated mobility responses for Retail. Left**, mean response. A 3D heatmap shows the average visitation reduction, with a clear monotonic decline as disease risk increases and policies become more restrictive. The accompanying 2D slices highlight marginal effects along each dimension. **Right**, variance response. A 3D heatmap depicts the sample variance of visitation changes, capturing the evolution of behavioral heterogeneity. The 2D projections indicate that variance decreases under higher risk and stricter policies, suggesting more uniformly reduced mobility across agents.

##### 3.2 Agent Number Comparison

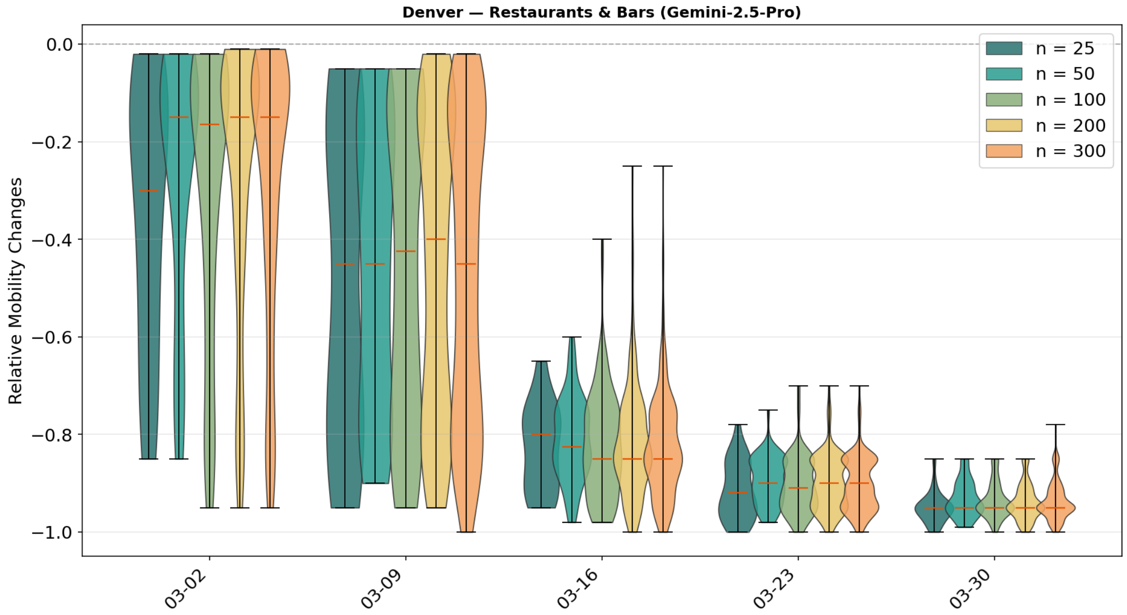

**Supplementary Figure 3: Sensitivity of Gemini-2.5-pro simulated mobility distributions to agent sample size in Denver.** The figure presents box plots showing the simulated distribution of relative mobility changes in Denver across five dates for POI type Restaurants & Bars. The model used in the figure is Gemini-2.5-pro.

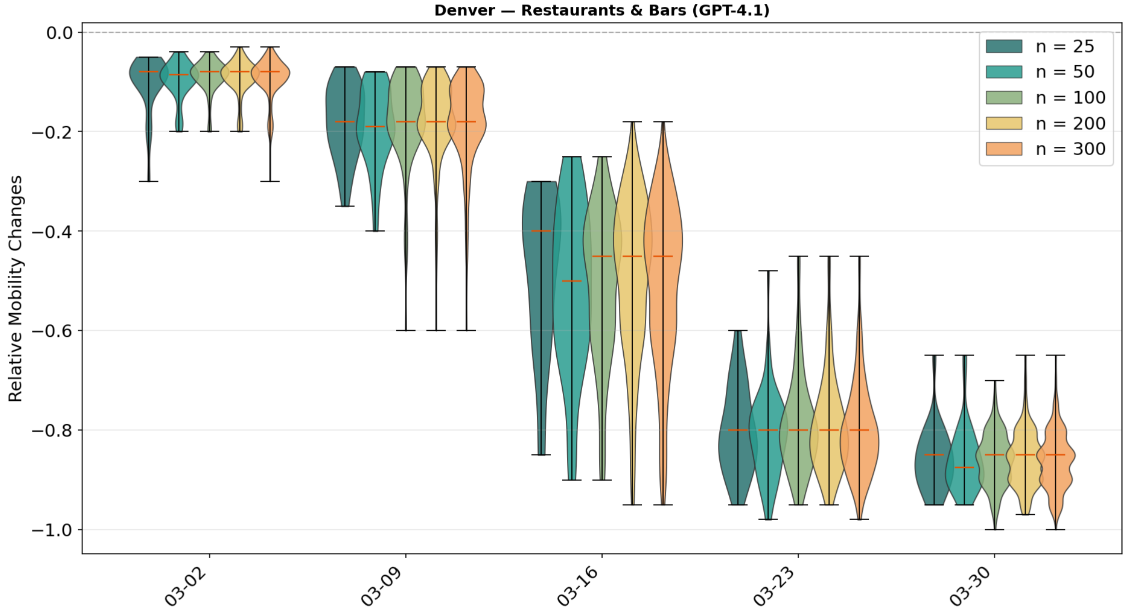

**Supplementary Figure 4: Sensitivity of GPT-4.1 simulated mobility distributions to agent sample size in Denver.** The figure presents box plots showing the simulated distribution of relative mobility changes in Denver across five dates for POI type Restaurants & Bars. The model used in the figure is GPT-4.1.

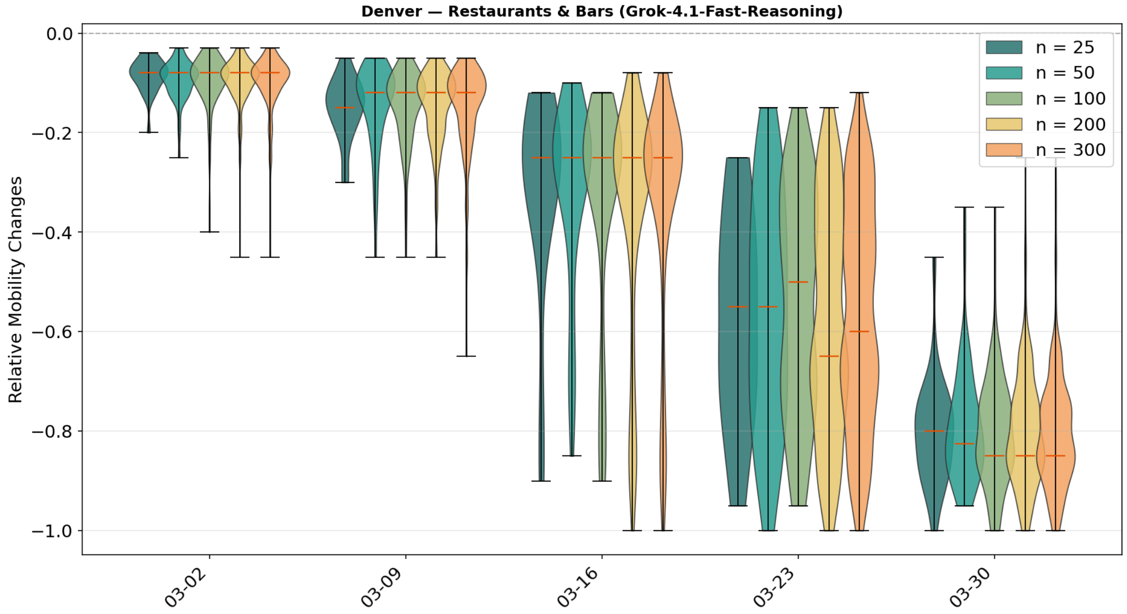

**Supplementary Figure 5: Sensitivity of Grok-4.1-Fast simulated mobility distributions to agent sample size in Denver.** The figure presents box plots showing the simulated distribution of relative mobility changes in Denver across five dates for POI type Restaurants & Bars. The model used in the figure is Grok-4.1-Fast.

##### 3.3 City Level Comparison across all Cities and all Models

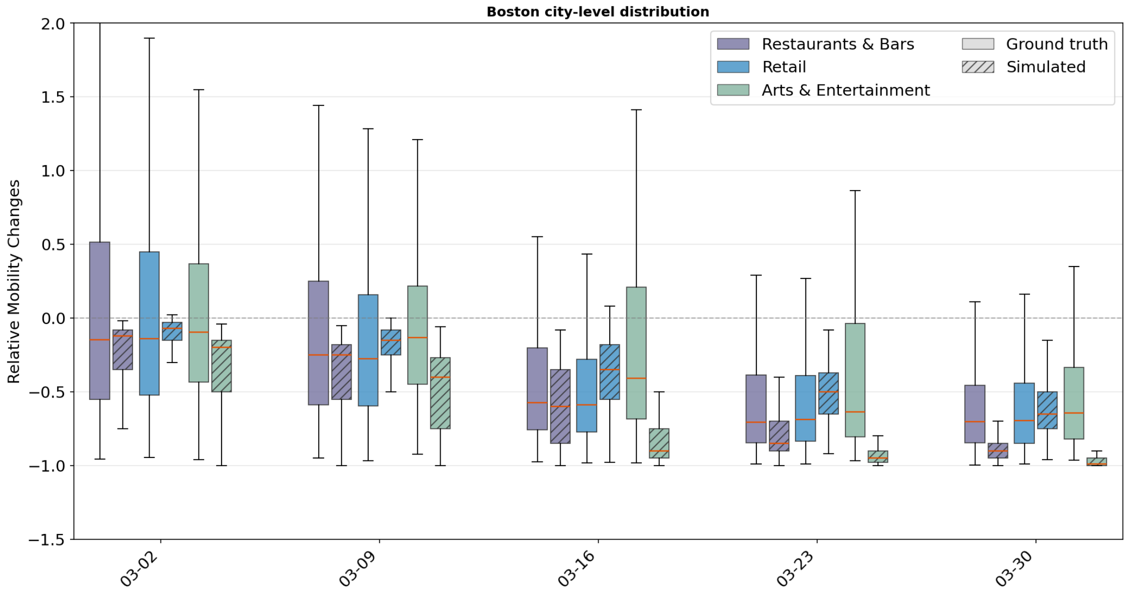

**Supplementary Figure 6: City-level distribution comparison of mobility responses in Boston.** The figure presents box plots showing the city-level distribution of visitation changes in Boston across five dates, comparing ground truth and simulated outputs for all three POI categories. The model used in the figure is the ensemble model.

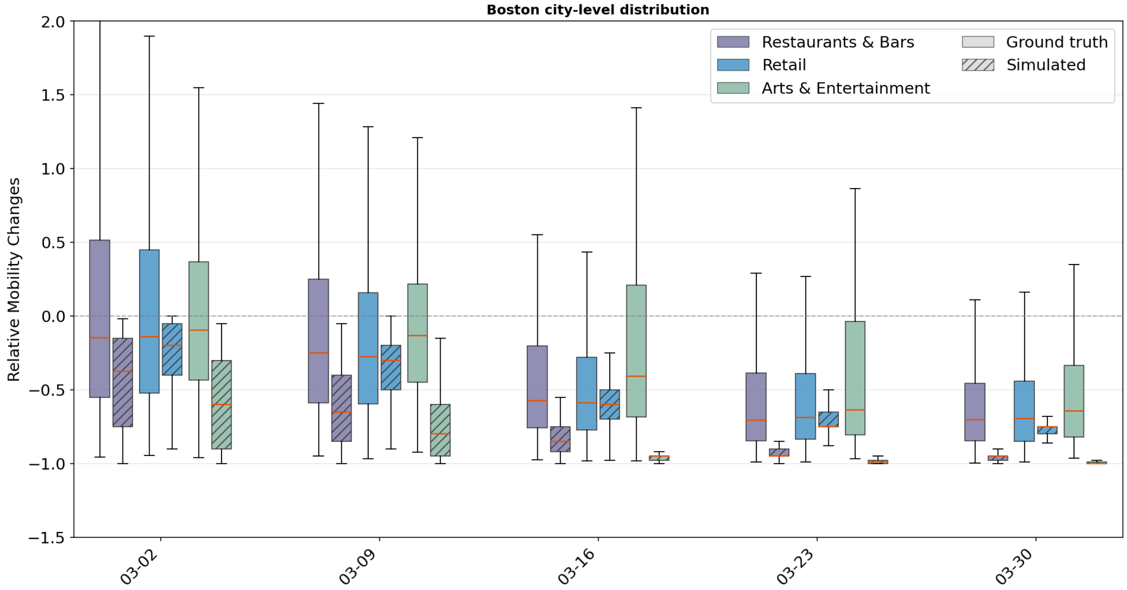

**Supplementary Figure 7: City-level distribution comparison of mobility responses in Boston.** The figure presents box plots showing the city-level distribution of visitation changes in Boston across five dates, comparing ground truth and simulated outputs for all three POI categories. The model used in the figure is the Gemini-2.5-pro model.

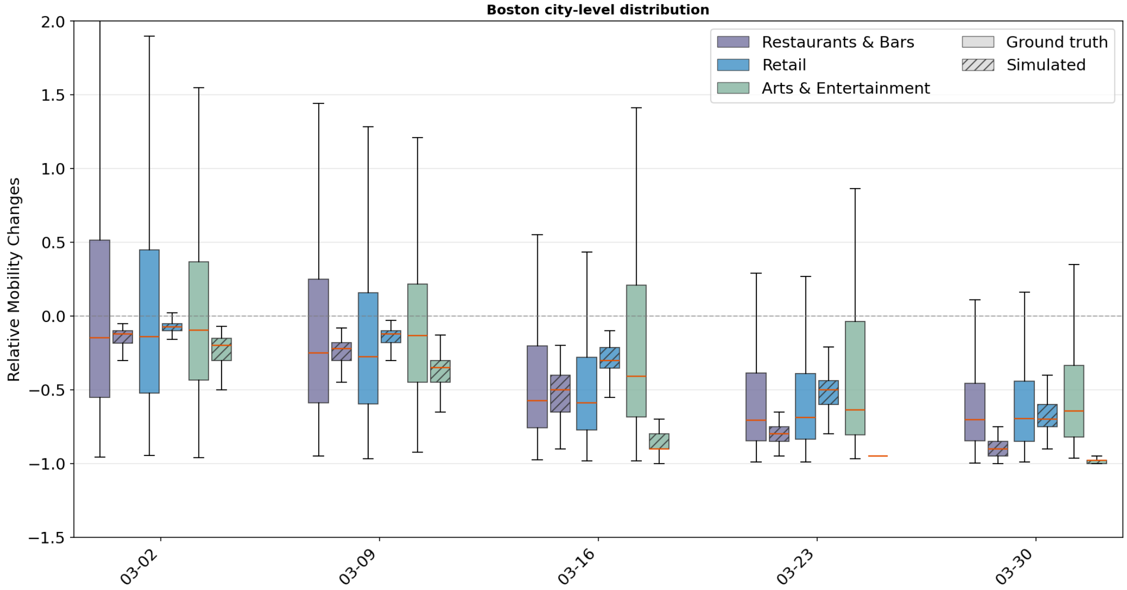

**Supplementary Figure 8: City-level distribution comparison of mobility responses in Boston.** The figure presents box plots showing the city-level distribution of visitation changes in Boston across five dates, comparing ground truth and simulated outputs for all three POI categories. The model used in the figure is the GPT-4.1 model.

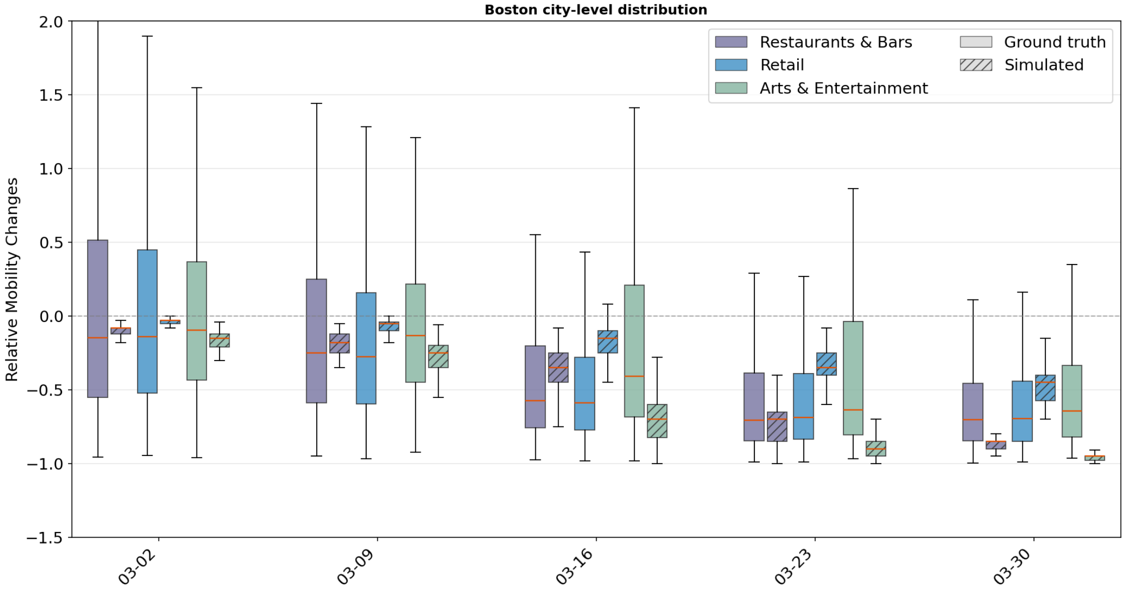

**Supplementary Figure 9: City-level distribution comparison of mobility responses in Boston.** The figure presents box plots showing the city-level distribution of visitation changes in Boston across five dates, comparing ground truth and simulated outputs for all three POI categories. The model used in the figure is the Grok-4.1-Fast model.

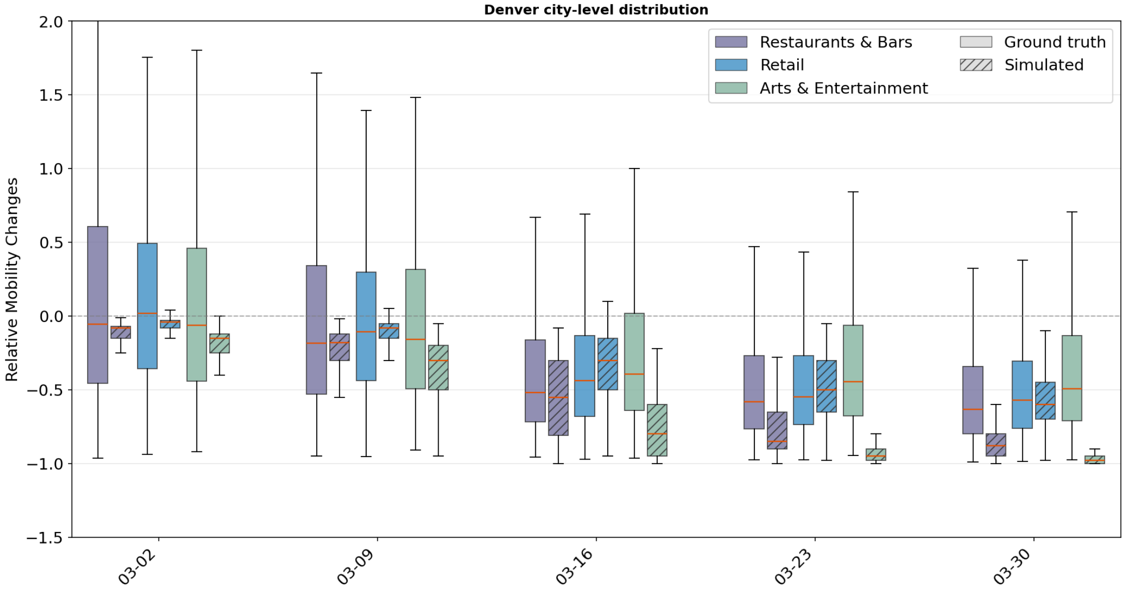

**Supplementary Figure 10: City-level distribution comparison of mobility responses in Denver.** The figure presents box plots showing the city-level distribution of visitation changes in Denver across five dates, comparing ground truth and simulated outputs for all three POI categories. The model used in the figure is the ensemble model.

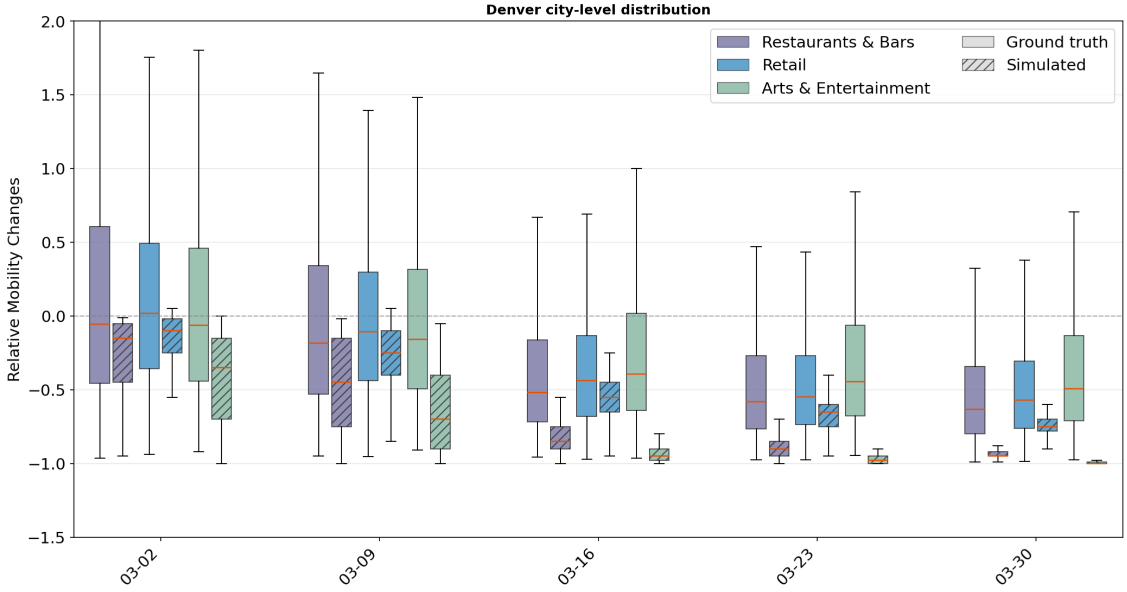

**Supplementary Figure 11: City-level distribution comparison of mobility responses in Denver.** The figure presents box plots showing the city-level distribution of visitation changes in Denver across five dates, comparing ground truth and simulated outputs for all three POI categories. The model used in the figure is the Gemini-2.5-pro model.

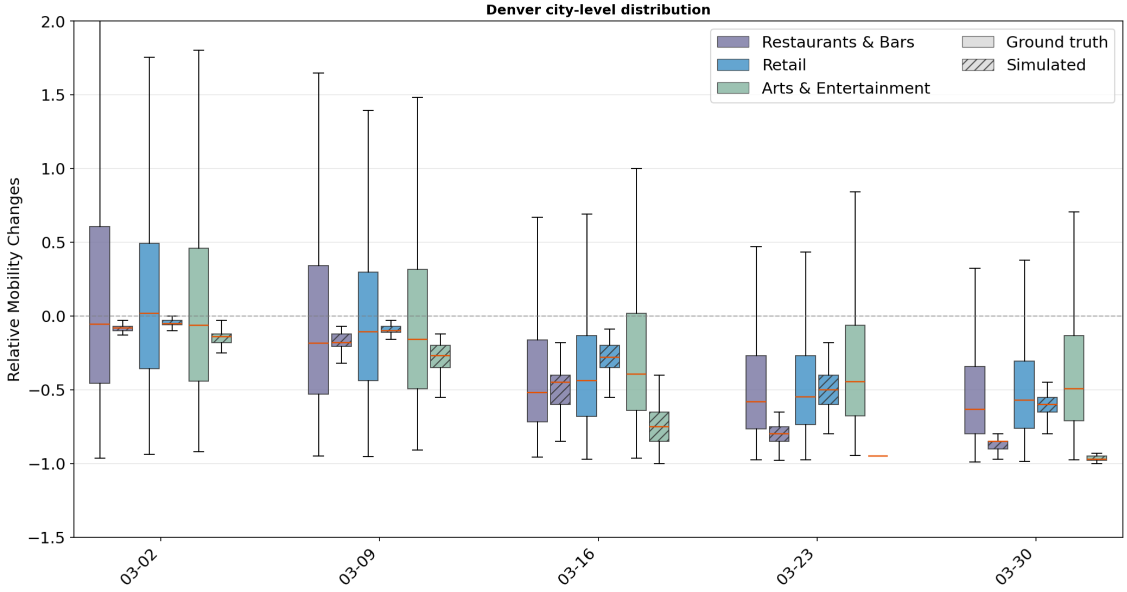

**Supplementary Figure 12: City-level distribution comparison of mobility responses in Denver.** The figure presents box plots showing the city-level distribution of visitation changes in Denver across five dates, comparing ground truth and simulated outputs for all three POI categories. The model used in the figure is the GPT-4.1 model.

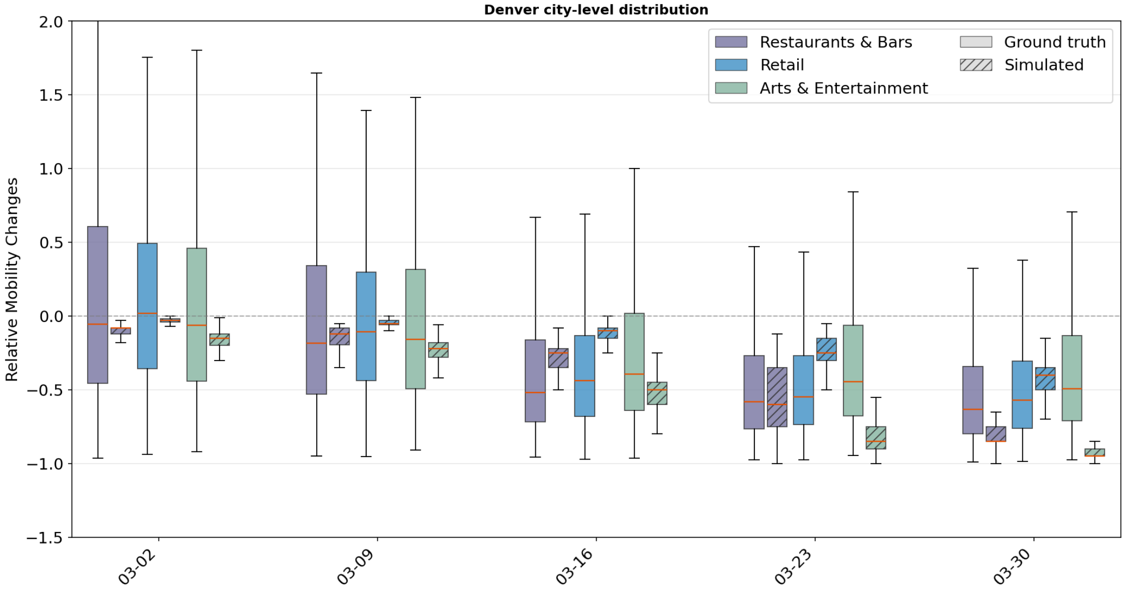

**Supplementary Figure 13: City-level distribution comparison of mobility responses in Denver.** The figure presents box plots showing the city-level distribution of visitation changes in Denver across five dates, comparing ground truth and simulated outputs for all three POI categories. The model used in the figure is the Grok-4.1-Fast model.

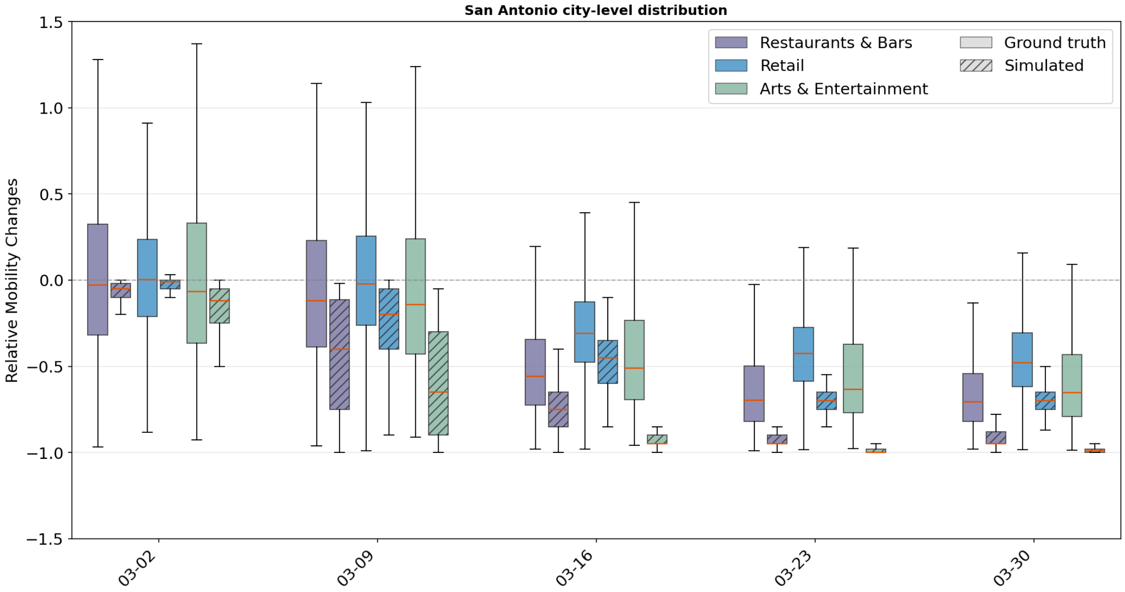

**Supplementary Figure 14: City-level distribution comparison of mobility responses in San Antonio.** The figure presents box plots showing the city-level distribution of visitation changes in San Antonio across five dates, comparing ground truth and simulated outputs for all three POI categories. The model used in the figure is the Gemini-2.5-pro model.

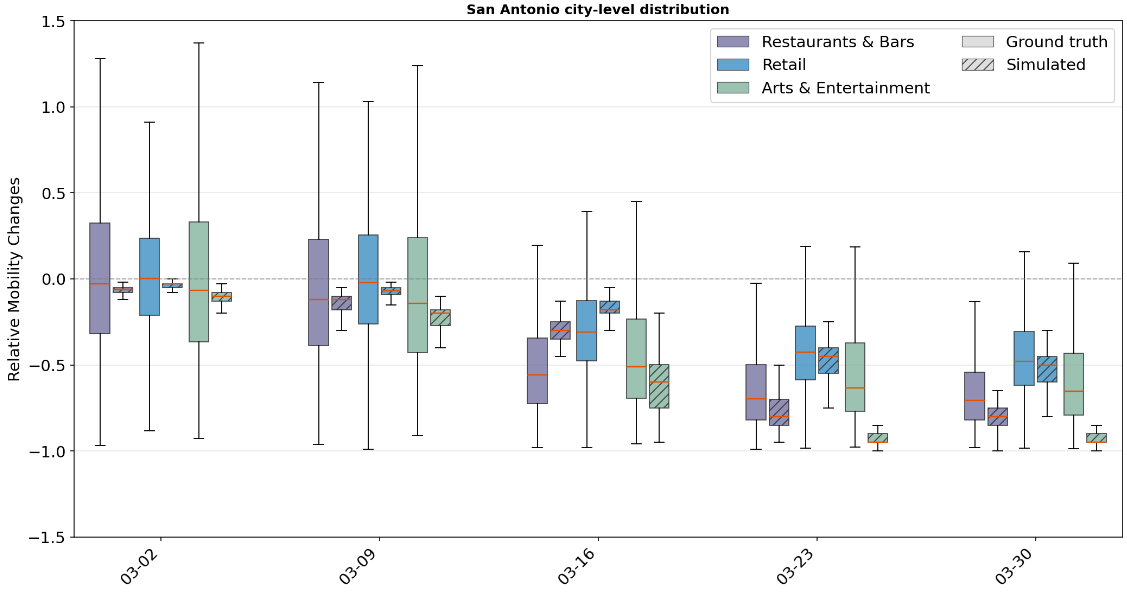

**Supplementary Figure 15: City-level distribution comparison of mobility responses in San Antonio.** The figure presents box plots showing the city-level distribution of visitation changes in San Antonio across five dates, comparing ground truth and simulated outputs for all three POI categories. The model used in the figure is the GPT-4.1 model.

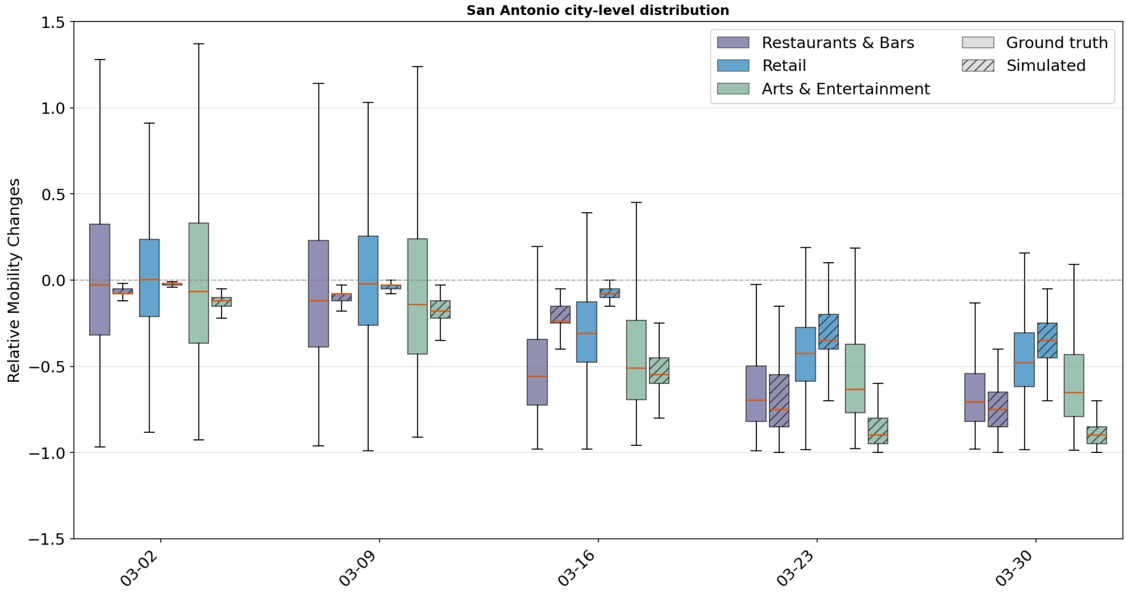

**Supplementary Figure 16: City-level distribution comparison of mobility responses in San Antonio.** The figure presents box plots showing the city-level distribution of visitation changes in San Antonio across five dates, comparing ground truth and simulated outputs for all three POI categories. The model used in the figure is the Grok-4.1-Fast model.

#### 3.4 Bar Chart for different Policies

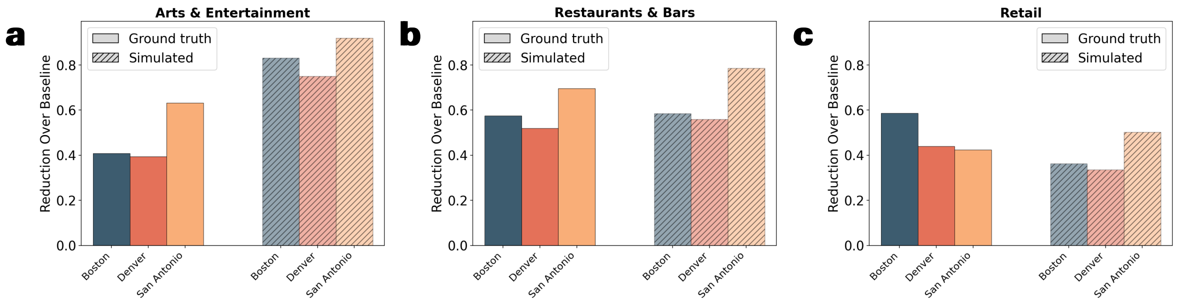

**Supplementary Figure 17: City-level comparison between observed and simulated mobility responses across POI categories. a–c**, Cumulative visitation reduction relative to baseline across Boston, Denver, and San Antonio for the same three POI categories: **a**, Arts & Entertainment, **b**, Restaurants & Bars, and **c**, Retail. Bars compare observed mobility reduction across the three cities on the week immediately following the policy "Protection of elderly people in LTCF" was enacted in each city.

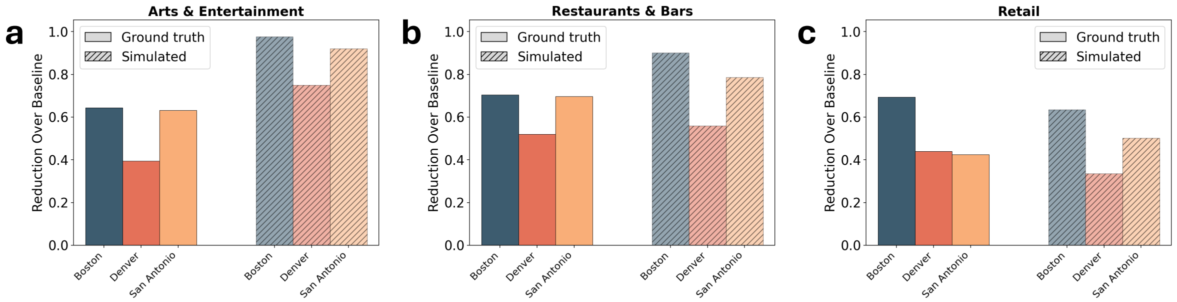

**Supplementary Figure 18: City-level comparison between observed and simulated mobility responses across POI categories. a–c**, Cumulative visitation reduction relative to baseline across Boston, Denver, and San Antonio for the same three POI categories: **a**, Arts & Entertainment, **b**, Restaurants & Bars, and **c**, Retail. Bars compare observed mobility reduction across the three cities on the week immediately following the policy "Workplace Closure" was enacted in each city.

#### 3.5 Prompt Sensitivity Analysis

To assess the stability of our model, we evaluated its performance under five different rephrased versions of the prompt. For each variant, we conducted experiment in Denver and compared the resulting distributions. As shown in the results, the box plots corresponding to all prompt versions across the three POIs exhibit highly similar distributions, indicating consistent behavior.

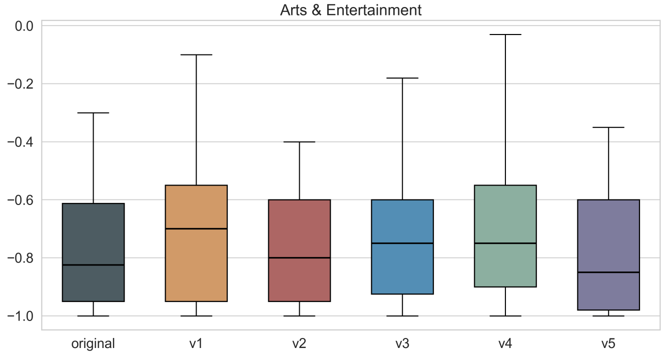

**Supplementary Figure 19: Prompt rephrasing stability analysis for Denver (03-16, Arts & Entertainment).** The figure presents box plots comparing simulation results across the original prompt and five rephrased variants for the Arts & Entertainment category on March 16 in Denver.

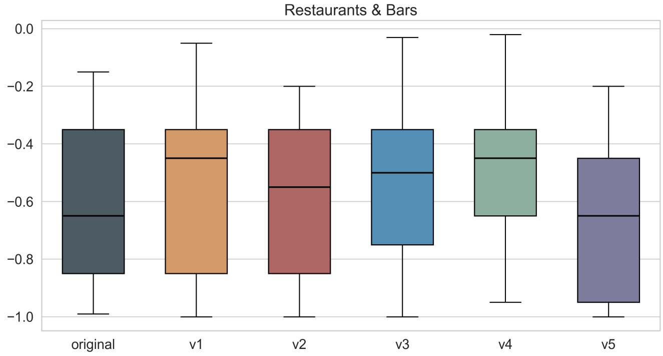

**Supplementary Figure 20: Prompt rephrasing stability analysis for Denver (03-16, Restaurants & Bars).** The figure presents box plots comparing simulation results across the original prompt and five rephrased variants for the Restaurants & Bars category on March 16 in Denver.

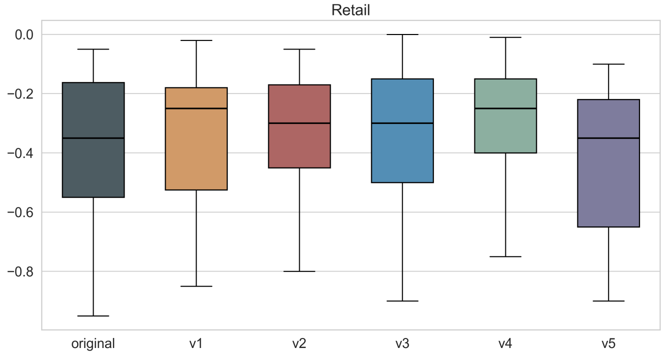

**Supplementary Figure 21: Prompt rephrasing stability analysis for Denver (03-16, Retail).** The figure presents box plots comparing simulation results across the original prompt and five rephrased variants for the Retail category on March 16 in Denver.
